# Trajectory of COVID-19 epidemic in Europe

**DOI:** 10.1101/2020.09.26.20202267

**Authors:** Marco Colombo, Joseph Mellor, Helen M Colhoun, M Gabriela M Gomes, Paul M McKeigue

**Author notes:** ARISING FROM S. Flaxman et al. Nature. https://www.nature.com/articles/s41586-020-2405-7 (2020).

## Abstract

The classic Susceptible-Infected-Recovered model formulated by Kermack and McKendrick [1] assumes that all individuals in the population are equally susceptible to infection. From fitting such a model to the trajectory of mortality from COVID-19 in 11 European countries up to 4 May 2020 Flaxman et al. concluded that “major non-pharmaceutical interventions – and lockdowns in particular – have had a large effect on reducing transmission” [2]. We show that relaxing the assumption of homogeneity to allow for individual variation in susceptibility or connectivity gives a model that has better fit to the data and more accurate 14-day forward prediction of mortality. Allowing for heterogeneity reduces the estimate of “counterfactual” deaths that would have occurred if there had been no interventions from 3.2 million to 262,000, implying that most of the slowing and reversal of COVID-19 mortality is explained by the build-up of herd immunity. The estimate of the herd immunity threshold depends on the value specified for the infection fatality ratio (IFR): a value of 0.3% for the IFR gives 15% for the average herd immunity threshold.

## Introduction

Kermack and McKendrick were careful to state in the abstract of their 1927 paper [1]:

> *In the present communication discussion will be limited to the case in which all members of the community are initially equally susceptible to the disease*

On this assumption the reproduction number ℛ_*t*_ at time *t* is (1−*p*_*t*_)ℛ_0_ where *p*_*t*_ is the proportion of the population that has been infected and is no longer susceptible. The herd immunity threshold *H* – the value of *p*_*t*_ at whichℛ_*t*_ = 1 – is thus 1 − 1*/* ℛ_0_. For natural infection, however, heterogeneity of susceptibility lowers the value of *H* [3]. If the distribution of susceptibility has a gamma distribution, in the expression above (1−*p*_*t*_) is replaced by (1−*p*_*t*_)^1+1*/α*^ where *α* is the shape parameter of the gamma distribution [4,5]. If connectivity, rather than susceptibility, has a gamma distribution, the exponent of (1−*p*_*t*_) is (1 + 2*/α*); models with heterogeneity of connectivity or heterogeneity of susceptibility are thus likelihood-equivalent. The classic model is thus a special case of this more general formulation, in which *α* = ∞ and the distribution of susceptibility or connectivity is a spike at 1.

## Methods

We compared a model that allows for heterogeneity of susceptibility with the original model that assumes homogeneity. The only change made to the original model was to replace the expression (1−*p*_*t*_) by the exponent form given above, with a half-Cauchy(0, 5) prior on the shape parameter *α*. Flaxman et al specified an average infection fatality ratio (IFR) of 1.1%, which is higher than most recent estimates. We repeated the model fitting with country-specific IFRs scaled by a factor of 0.275 to equate the average IFR to a recent estimate [6] of 0.30%. For sensitivity analyses, we repeated the comparison of homogeneity and heterogeneity models after varying other modelling assumptions:

1. Replacing the sparsity-enforcing gamma priors [7] on the effects of interventions with half-normal priors.
2. Removing country-specific effects, specified in the original model for the last intervention only.
3. Fade-in of effects of intervention with half-life of one day, by scaling the indicator variable for each intervention by a factor of (1−e^−0.7*d*^), where *d* is the number of days since the start of the intervention.

## Results

Table 1 compares the homogeneity and heterogeneity models. In comparison with the homogeneity model, the heterogeneity model has better fit to the data (deviance reduced by 23 units at the cost of one extra parameter) and better 14-day forward predictive performance (mean squared error reduced by 59%). When other modelling assumptions are varied, the heterogeneity model still has better fit than the homogeneity model (Supplementary Table).

**Table 1.** Comparison of original model with model that allows heterogeneity, fitted to 128925 deaths from COVID-19 in 11 European countries. Ranges are across countries

|  | IFR 1.08% (original model) |  | IFR 0.3% |  |
| --- | --- | --- | --- | --- |
|  | Homogeneity | Heterogeneity | Homogeneity | Heterogeneity |
| Deviance $D$ ($2 \times$<br>nat log units) | 5628.4 | 5605.1 | 5624.0 | 5603.9 |
| Effective number of<br>parameters $p_D$ | 27.2 | 28.3 | 26.7 | 28.3 |
| Deviance<br>Information<br>Criterion ($D + p_D$) | 5655.5 | 5633.5 | 5650.7 | 5632.2 |
| Basic reproduction<br>number $\mathcal{R}_0$ | 3.9 (2.7, 5.2) | 3.9 (2.9, 5.1) | 4.0 (2.8, 5.4) | 4.0 (3.0, 5.1) |
| Shape parameter of<br>susceptibility<br>distribution $\alpha$ | ∞ | 0.03 | ∞ | 0.15 |
| Herd immunity<br>threshold $H$ | 0.728 (0.628,<br>0.805) | 0.039 (0.031,<br>0.047) | 0.735 (0.640,<br>0.812) | 0.154 (0.125,<br>0.181) |
| Total infections | 13,382,481 | 12,610,313 | 48,351,305 | 45,834,605 |
| Expected deaths | 128,797 | 129,664 | 129,132 | 129,834 |
| Expected<br>counterfactual<br>deaths | 3,238,848 | 262,102 | 1,017,185 | 276,624 |
| Mean squared error<br>of 14-day forward<br>prediction | 528,243.8 | 216,655.3 | 417,011.2 | 219,052.8 |

With the original mean IFR setting of 1.1%, the estimated number of infections is about 13 million out of a total population of 374 million. Under a homogeneity model, the early removal of susceptible individuals can slow the growth of the epidemic only slightly, so the counterfactual estimate of deaths that would have occurred without intervention is 3.2 million. Under the heterogeneity model, the growth of the epidemic is slowed and reversed by the build-up of herd immunity, so that the counterfactual estimate of deaths is only 262,000. Rescaling the IFRs to an average of 0.3% increases the imputed number of infections approximately by the inverse of the scaling factor in both models. With this setting of the IFR, the herd immunity threshold estimate (average, inter-country range) was 0.154 (0.125, 0.181) under the heterogeneity model, compared with 0.735 (0.640, 0.812) under the homogeneity model.

Fig 1 compares the two fitted models for the United Kingdom with the original IFR setting. Under the heterogeneity model, the stepwise effect of lockdown on the imputed daily number of infections is smaller than under the homogeneity model, and the trajectory of the reproduction number ℛ_*t*_ up to the date of lockdown approximates a smooth reverse sigmoid curve.

**Fig 1.**
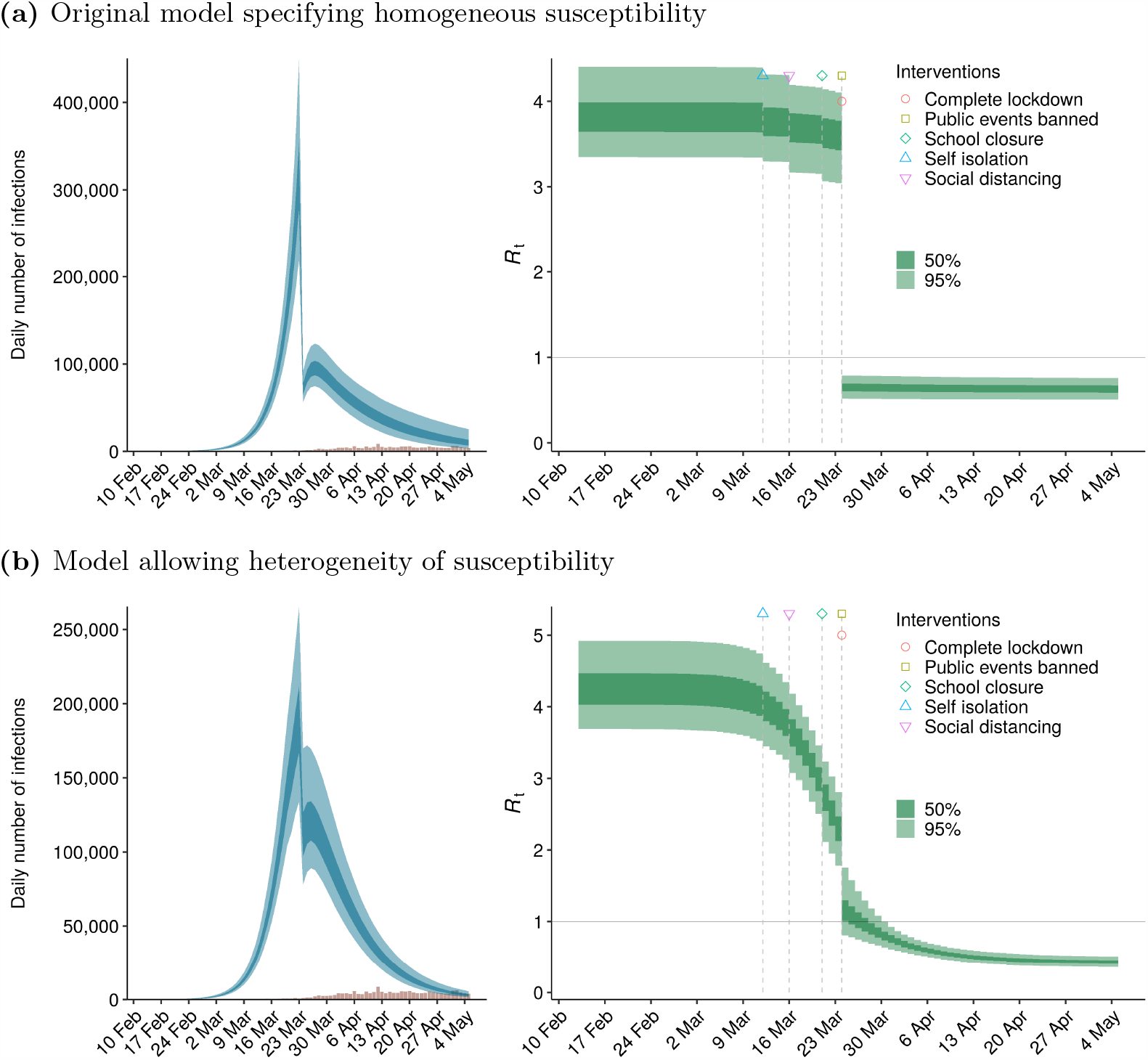
Comparison of homogeneity and heterogeneity models for the United Kingdom: imputed epidemic curves and trajectory of reproduction number *R*_*t*_

## Discussion

Models that allow for heterogeneity favour build-up of herd immunity rather than non-pharmaceutical interventions as the main factor underlying the early slowing and reversal of the COVID-19 epidemic in Europe. This is consistent with observations that epidemic curves in many countries reached a peak less than two months after the first few severe cases appeared [8,9]. With this dataset it is not possible to distinguish the relative contributions of heterogeneity of connectivity, heterogeneity of susceptibility, or any other process that could have generated a smooth downward trajectory in R_*t*_ over about one month in each of the 11 European countries studied.

Because the model is fitted to observed deaths, the estimates of cumulative numbers infected and herd immunity threshold depend on the values pre-specified for infection fatality ratios. Specifying an average infection fatality ratio of 0.3% gives an estimated herd immunity threshold of 15%. Whatever value is specified for the infection fatality ratio, a model that allows for heterogeneity has better fit to the data than the homogeneity model and supports herd immunity as the main factor underlying the reversal of the epidemic.

One objection that has been raised to estimates that herd immunity thresholds for COVID-19 are less than 20% is that far higher infection rates have been reached in local hotspots such as Manaus [10]. However country-level herd immunity thresholds as estimated here are not likely to be homogeneous over every locality. In hotspots where the basic reproduction number ℛ_0_ is higher than the population average, the herd immunity threshold and overshoot of this threshold will be correspondingly higher, with or without heterogeneity.

The release of the modelling code and dataset used by Flaxman et al is a valuable contribution to transparent evaluation of infectious disease modelling. However once the unrealistic assumption of no individual variation in susceptibility or connectivity is relaxed, the model does not support their estimate that lockdown reduced the case reproduction number *R* by 81% or that more than three million deaths were averted by non-pharmaceutical interventions.

## Data Availability

All data and modelling code are freely available at the URL given below.

https://github.com/mcol/covid19model

## Author contributions

PM, GG and HC conceived the study. MC and JM undertook the modelling and computational work. All authors contributed to drafting this note.

## Code availability

Code used in this reanalysis is available on [GitHub](https://github.com/mcol/covid19model-reanalysis).

## Supplementary table: sensitivity to modelling assumptions

**Table 2.** Comparison of homogeneity and heterogeneity models: sensitivity to modelling assumptions

|  | Homogeneity | Heterogeneity |
| --- | --- | --- |
| **Half-normal priors on effects of interventions** |  |  |
| Deviance ($2 \times \text{nat log units}$) | 5629.9 | 5604.7 |
| Effective number of parameters $p_D$ | 26.7 | 28.5 |
| Deviance Information Criterion | 5656.6 | 5633.2 |
| Basic reproduction number $R_0$ | 4.3 (3.0, 5.9) | 4.3 (3.3, 5.4) |
| Shape parameter $\alpha$ | ∞ | 0.03 |
| Herd immunity threshold $H$ | 0.754 (0.664, 0.828) | 0.043 (0.035, 0.050) |
| Total infections | 13,421,323 | 12,611,749 |
| Expected deaths | 128,061 | 128,972 |
| Expected counterfactual deaths | 3,525,987 | 287,545 |
| **No country-specific effects** |  |  |
| Deviance ($2 \times \text{nat log units}$) | 5672.8 | 5650.8 |
| Effective number of parameters $p_D$ | 21.8 | 23.8 |
| Deviance Information Criterion | 5694.6 | 5674.6 |
| Basic reproduction number $R_0$ | 4.2 (2.2, 5.9) | 3.9 (2.4, 6.7) |
| Shape parameter $\alpha$ | ∞ | 0.07 |
| Herd immunity threshold $H$ | 0.745 (0.542, 0.828) | 0.078 (0.053, 0.110) |
| Total infections | 13,761,302 | 12,966,248 |
| Expected deaths | 129,128 | 129,001 |
| Expected counterfactual deaths | 3,389,856 | 488,031 |
| **Fade-in of effects of interventions** |  |  |
| Deviance ($2 \times \text{nat log units}$) | 5625.2 | 5605.7 |
| Effective number of parameters $p_D$ | 27.0 | 28.3 |
| Deviance Information Criterion | 5652.2 | 5633.9 |
| Basic reproduction number $R_0$ | 3.8 (2.7, 5.1) | 3.9 (2.9, 5.0) |
| Shape parameter $\alpha$ | ∞ | 0.03 |
| Herd immunity threshold $H$ | 0.722 (0.626, 0.799) | 0.041 (0.033, 0.049) |
| Total infections | 13,309,353 | 12,593,876 |
| Expected deaths | 129,471 | 129,897 |
| Expected counterfactual deaths | 3,186,110 | 273,675 |

